# Impact of COVID-19 on the national development of countries: implications for the public health

**DOI:** 10.1101/2022.10.24.22281450

**Authors:** Olha Kuzmenko, Serhiy Lyeonov, Nataliia Letunovska, Mariya Kashcha, Wadim Strielkowski

## Abstract

The article focuses on measuring the fluctuations in countries’ development as a result of the COVID-19 pandemic. The obtained measures make it possible to predict the extent of the impact of risks to public health on the economy, financial-budgetary, political-institutional development of states in the future, as well as the social determinants of public health. This assessment represents a new paradigm that makes it possible to effectively evaluate the manifestations of the consequences of COVID-19 and to identify the relevant determinants of the lack of resilience of the medical and social security systems to the coronavirus pandemic around the world. We picked the determinant of national development indicators of the 59 countries in order to measure the fluctuations in their economic development. In addition, we applied the binary response model for identifying the economic, financial-budgetary, and political-institutional development change with the happiness index of the countries being the dependent variable. The analysis of our empirical model made it possible for us to conclude that economic and financial-budgetary components have significantly increased the influence on well-being during the COVID-19 pandemic. In contrast, we observed the decrease in the impact of political and institutional indicators during the same period.

## 1. Introduction

Despite the duration of the emergency state in the society, the COVID-19 pandemic that officially started in March 2020 is still responsible for the unhealthy environment in the social and economic aspects of life all around the world (Hardy et al. 2021; Kumar et al., 2021; Strielkowski et al. 2022). Scientists use the term “pandemic” to refer to the extraordinary efforts of the society in the fight against a dangerous virus that has changed the way people are living today and that induced the anti-epidemic policies that needed to be developed and implemented by many governments around the world (Bernacer et al. 2021; Sandset and Villadsen, 2022). Concerns over the existence of a threat to the public health forces researchers and scientists at all levels to permanently engage in the search for the ways to overcome the pandemic’s negative medical and financial consequences and, in the current conditions, to level the possible impacts of the pandemic over time. In the summer of 2022, according to the World Health Organization (COVID-19 weekly, 2022), there was an increase in regional cases of various strains of the coronavirus in the Eastern Mediterranean Region (+29%), the South-East Asia Region (+20%), the European Region (+15%), the Western Pacific Region (+4%). And even these trends need to be interpreted with caution, as some countries are gradually changing their strategies for identifying cases of COVID-19 which leads to the lower totals.

As of August 2022, at least 6.4 million people have died from the virus, and many more continue to suffer from the adverse long-term health effects of the infection (Coronavirus, 2022). At the same time, measures to combat the spread of the virus caused and continue to cause damage to the world’s economy. Many countries risk being left behind as the developed countries are recovering from the pandemic. They may spend significant financial and time resources to recover from the crisis caused by the COVID-19. Ultimately, they may make little progress towards the Sustainable Development Goals (SDGs). The current situation is exceptional and requires decisive action by the international community to counter the risks. Given these challenges, it is essential to consider what lessons their past and projected trajectories can provide to inform how best to lay the foundations for a sustainable recovery from the shocks of COVID-19. Additionally, it appears to be relevant to measure the fluctuations in the development of the countries as a result of the COVID-19 pandemic, which will make it possible to predict the extent of the impact of risks to public health on the economy, financial-budgetary and political-institutional development of the states in the future. This very assessment represents a new paradigm that makes it possible to more effectively evaluate the manifestations of the consequences of COVID-19 and to identify the relevant determinants of the lack of resilience of the world’s population’s medical and social security system to the virus.

The paper is organized as follows: Section 2 provides a comprehensive literature review. Section 3 explains the methodology used in our study. Section 4 outlines the empirical results that includes the selection of relevant indicators, construction of integral indicators of manifestation of the consequences of COVID-19, and the implementation of probit/logit modeling of the manifestation of the consequences of the COVID-19 pandemic. Finally, section 5 provides the conclusions and outlines the implementations of the study.

## 2. Literature Review

Studies in which scientists try to measure the degree of transformation of the economic, social, political, institutional, financial-budgetary and other spheres of society in connection with the spread of the COVID-19 pandemic are shared among the world scientific community (Kuzmenko et al., 2021; Strielkowski et al. 2021). The studies which represent the particular interest with regard to this problem are the ones that are resolved with the application of the econometric tools and models. Kitenge (2020) theoretically proved the absence of a relationship between the vulnerability to COVID-19 and the income of a person, using probit/logit modeling. Huterska et al. (2021) used logistic regression for modeling the impact on the socio-economic life of the population during the pandemic. Vu and Ho (2022) used a similar toolkit to determine credit availability for persons engaged in informal work during quarantine restrictions, while Al-Ahmadi and Kasztelnik (2021) investigated the labor market fluctuations as one of the essential components of an efficient economy (see Kurian, 2021) They also draw attention in their study to the negative consequences of the pandemic for achieving the Sustainable Development Goals (SDG), even considering the revision of priorities for their achievement. It should also be noted the works whose authors explore ways to overcome the negative economic consequences of the pandemic. For example, Ray (2021) concluded that vaccination and mass immunization of the population is a powerful tool for combating the disease and, as a result, a path to stability in the state and establishment of global sustainable development. Some authors (see Ed. Fernando Alonso Ojeda Castro, 2021; or Law, 2021) see the possibility of overcoming the negative consequences of the pandemic through the development of innovative policies, the creation of virtual banks, and the implementation of effective measures to achieve cyber security at the global level. Scientists (Fast, 2021) consider various scenarios for exiting the economic crisis, relying on the scientific works of Keynes and Hayek and world experience, analyzing the speed of recovery from previous recessions. Biewendt et. al. (2021) emphasize that the quarantine restrictions have negatively affected the business sphere, so the authors draw attention to the need for immediate transformations in management and elimination of the lack of motivation among workers. (Sardak, 2018) determined even before the pandemic that social problems and the consequences of risks, in particular in the field of health, cause significant changes in the overall development of the system. Many studies by scientists draw attention to transformations due to the pandemic in various spheres of social life (see e.g. Kuzmenko et al., 2020; Vasilyeva et al., 2021). Moskovicz (2021) does the same for the entrepreneurship and draws attention to the positive changes in the financing of university startups, which have a significant impact on the development of innovative activities. Keliuotytė-Staniulėnienė and Daunaravičiūtė (2021) focus on the global green bond market, while Hinrichs and Bundtzen (2021) tackle the insurance activity, in particular, the role and new opportunities of the insurance agent. Bouchetara et al. (2020) do the same for the banking sector, through macroprudential policy instruments, while Albliwi and Alsolami (2021) do in the development of electronic commerce in the world. Lyulyov et al. (2021) and Khvostina et. al. (2021) do the same in ecology, through the construction of an integral risk indicator, while Koibichuk et. al. (2021) tackle the development of cyber fraud and the need to develop innovative technologies to combat them. Vasudevan and Aslan (2021) focus on the field of services, and the impact of marketing technologies on its development. While Hanulakova et al. (2021) do this in the medical field, which has probably undergone the most challenges and changes. Samusevych et al. (2021a) in education, due to the possibility of loss of knowledge. In Antonyuk et al. (2021), the authors note that the pandemic has significantly affected business conditions, changing the priorities and principles of the economy of almost every country in the world. Their research aimed to analyze the impact of quarantine measures and the pandemic on further business development to ensure sustainable development. Other authors (Vasylieva et al., 2021b; Samusevych et al., 2021b; Tiutiunyk et al., 2021) analyzed the consequences of COVID-19 in important spheres of public functioning, namely taxes, informatization, digitalization etc. Among the negative impacts of the pandemic, the stratification of society and a significant psycho-emotional burden, which threatens socio-economic development, have been identified. Boronos et al. (2020) pointed out at the issue of ensuring financial security and business resilience to the impact of COVID-19. Thus, the negative consequences of the pandemic crisis manifested themselves in the deterioration of the financial results of business entities and the financial sector as a whole (Vasylieva et al., 2021a; Moskalenko et al., 2022), especially in the industrial sphere, in the transport, hotel, and restaurant business. (Smiianov et al., 2020a) Their study formed a methodological basis for assessing socio-economic trends in the functioning of the labor market in the health care field in the context of prevention and countermeasures against epidemic threats. In Smiianov et al. (2020b), Kuznyetsova et al., (2021), the authors developed a methodology to test the hypothesis of a link between the consequences of pandemic quarantine and public health and economic growth and country security. Romanello et al. (2021) or Kwilinski et al. (2022) analyzed the existence of a relationship between the state of the country’s energy sector and key indicators of population health, particularly resilience to the impact of pandemic threats.

Therefore, among the world’s scientists, researching the transformation of various spheres of life in connection with the emergence of COVID-19 is relevant because there are many publications on various topics, which are united by one question – the impact of the pandemic. Also, the methods of constructing integral indicators, applying regression-correlation analysis, and logit/probit modeling are widespread. However, the combination of integral assessment of the manifestation of the consequences of COVID-19 in the economic, social, political, institutional, financial, and budgetary spheres of society through the use of additive-multiplicative convolutions and logit/probit modeling is insufficiently applied.

## 3. Methodology

In our paper, we have selected the 59 countries of the world including the following ones: Australia, Austria, Belgium, Bosnia and Herzegovina, Brazil, Canada, Switzerland, Chile, China, Cyprus, Czech Republic, Germany, Denmark, Spain, Estonia, Ethiopia, Finland, France, United Kingdom, Georgia, Greece, Honduras, Croatia, Hungary, Indonesia, India, Ireland, Iceland, Israel, Italy, Japan, Kazakhstan, Lithuania, Luxembourg, Latvia, Moldova, North Macedonia, Malta, Montenegro, Malaysia, Nigeria, Netherlands, Norway, Panama, Peru, Philippines, Poland, Portugal, Romania, Russian Federation, Serbia, Slovak Republic, Slovenia, Sweden, Thailand, Turkey, Tanzania, Ukraine and the United States. The time range was 2017-2019 for modeling changes in the economic, budget-financial and political-institutional development of countries before the COVID-19 pandemic and 2020 for modeling the manifestation of the consequences of the pandemic. The sites such as: statista.com, theglobaleconomy.com, and ec.europa.eu became the information base for this research.

In total, nine determinants were chosen to measure fluctuations in the economic development of the countries of the world due to the pandemic: exports of goods and services (% of GDP), Imports of goods and services (% of GDP), GDP growth (annual %), Inflation, consumer prices (annual %), Personal remittances, received (% of GDP), Gross savings (% of GDP), GNI per capita, Atlas method (current US$) Household consumption, billion U.S. dollars, Unemployment rate, %.; for budget and financial 7: Bank capital to assets ratio (%), Banking system z-scores, index points, Bank non-performing loans to total gross loans (%), Commercial bank branches (per 100,000 adults), Total reserves (includes gold, current US$), General government debt (% of GDP), Capital investments (% of GDP).; political and institutional - Corruption Perceptions Index, Democratic performance numeric, Property Rights Index, Voice and accountability, Political stability, and Government effectiveness.

## 4. Empirical results

### 4.1. Indicators of economic, budgetary, financial, political, and institutional development

In order to reduce the data in the set of determinants indicating the financial and budgetary development of countries, the Statistica Portable application program package was used using the Multivariate Exploratory Techniques/Principal Components and Classification Analysis toolkit, which makes it possible to classify variables by degree of relevance by diagonalizing the correlation matrix. For a set of financial and budgetary determinants, a stony scree graph was constructed (Fig. 1), which, according to Kettel’s criterion, clearly demonstrates the number of factors that must be included in the study to ensure maximum variation in space with a smaller number of variables (Polyakov et al., 2019). Tables 1–3 also show the percentage of variance explained by each factor, the cumulative eigenvalue of the corresponding characteristic, and the variance.

**Figure 1.**
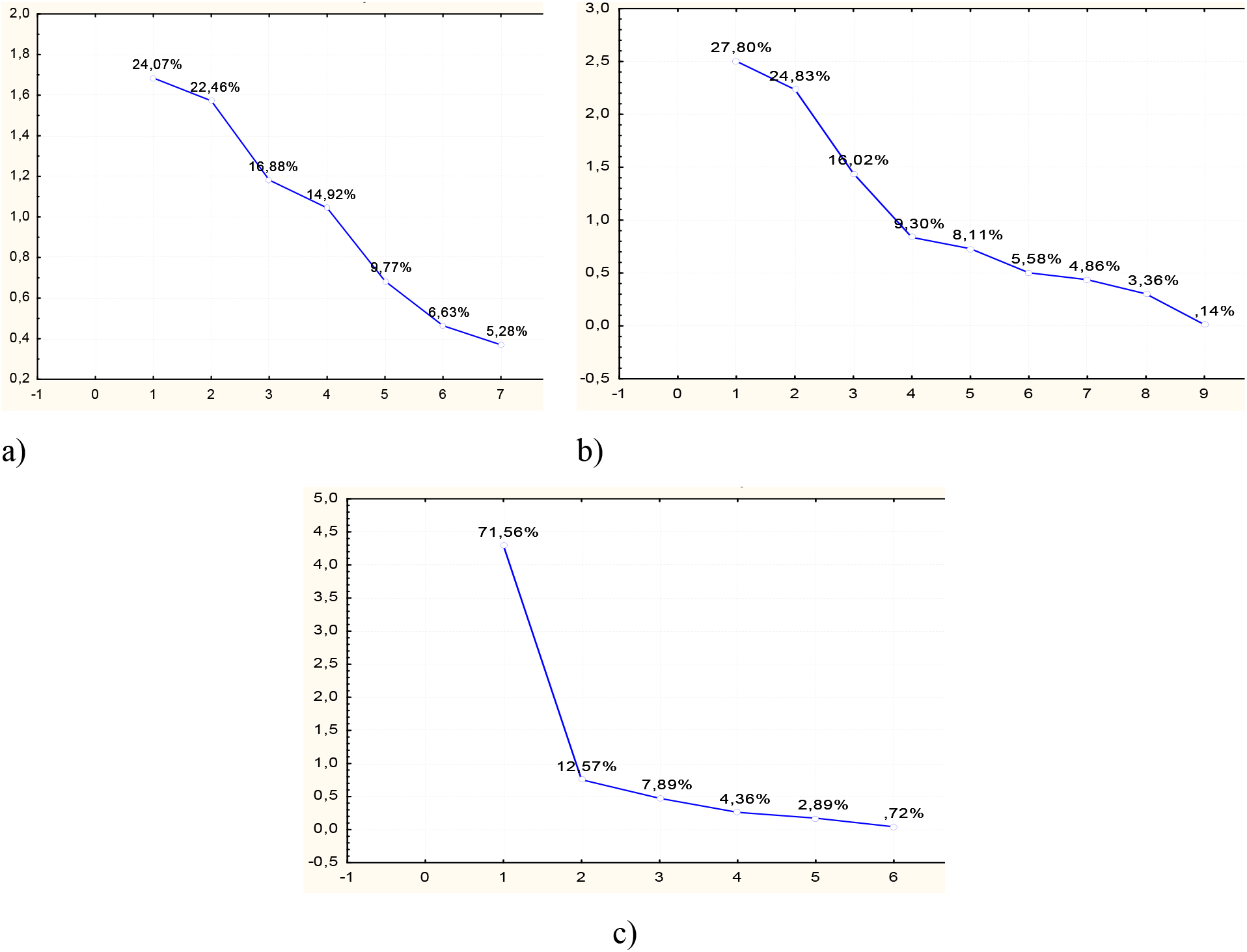
Stony scree for determinants (a – financial-budgetary, b – economic, c – political) Source: own results

**Table 1.** Eigenvalues of indicators of financial and budgetary development

|  | Eigenvalue | % Total – variance | Cumulative – Eigenvalue | Cumulative – % |
| --- | --- | --- | --- | --- |
| 1 | 1.684560 | 24.06514 | 1.684560 | 24.0651 |
| 2 | 1.572152 | 22.45931 | 3.256712 | 46.5245 |
| 3 | 1.181674 | 16.88105 | 4.438385 | 63.4055 |
| 4 | 1.044377 | 14.91967 | 5.482762 | 78.3252 |
| 5 | 0.683740 | 9.76772 | 6.166502 | 88.0929 |
| 6 | 0.463961 | 6.62801 | 6.630463 | 94.7209 |
| 7 | 0.369537 | 5.27910 | 7.000000 | 100.0000 |

Source: own results

**Table 2.** Eigenvalues of indicators of economic development

|  | Eigenvalue | % Total – variance | Cumulative – Eigenvalue | Cumulative – % |
| --- | --- | --- | --- | --- |
| 1 | 2.502182 | 27.80203 | 2.502182 | 27.8020 |
| 2 | 2.234374 | 24.82638 | 4.736556 | 52.6284 |
| 3 | 1.441463 | 16.01625 | 6.178019 | 68.6447 |
| 4 | 0.837182 | 9.30202 | 7.015201 | 77.9467 |
| 5 | 0.730051 | 8.11168 | 7.745252 | 86.0584 |
| 6 | 0.502182 | 5.57980 | 8.247435 | 91.6382 |
| 7 | 0.437490 | 4.86100 | 8.684925 | 96.4992 |
| 8 | 0.302765 | 3.36406 | 8.987690 | 99.8632 |
| 9 | 0.012310 | 0.13678 | 9.000000 | 100.0000 |

Source: own results

**Table 3.** Eigenvalues of indicators of political and institutional development

|  | Eigenvalue | % Total – variance | Cumulative – Eigenvalue | Cumulative – % |
| --- | --- | --- | --- | --- |
| 1 | 4.256198 | 60.80283 | 4.256198 | 60.8028 |
| 2 | 1.097807 | 15.68296 | 5.354005 | 76.4858 |
| 3 | 0.669839 | 9.56913 | 6.023845 | 86.0549 |
| 4 | 0.580194 | 8.28849 | 6.604039 | 94.3434 |
| 5 | 0.243484 | 3.47834 | 6.847523 | 97.8218 |
| 6 | 0.093369 | 1.33384 | 6.940892 | 99.1556 |
| 7 | 0.059108 | 0.84440 | 7.000000 | 100.0000 |

Source: own results

The analysis of the schedule allows us to conclude that for the next stage of the research, it is necessary to include the number of factors that provide a cumulative variation at the level of at least 75% and have an intrinsic value greater than one, that is, for the budget and financial determinant, these are four factors (78.3%), for economic – 4 factors (77.9%) and political and institutional – 2 factors (84.1%).

For determining the set of relevant determinants and their priority for inclusion in the study, a table of eigenvalues of the correlation matrix (tables 4–6) was built, including the weight of each variable’s contribution to each factor to filter out less relevant indicators.

**Table 4.** The contribution of financial and budgetary variables to each factor

| | Factor 1 | Factor 2 | Factor 3 | Factor 4 | $\sum_1^n \omega_j \cdot f_{ij}$ |
| --- | --- | --- | --- | --- | --- |
| Bank non-performing loans | 0.17829 | 0.11522 | 0.01252 | 0.33751 | 10.150 |
| Bank capital to assets ratio | 0.02095 | 0.24861 | 0.18064 | 0.06789 | 9.689 |
| Commercial bank branches | 0.10356 | 0.11263 | 0.12005 | 0.36838 | 10.455 |
| Total reserves | 0.08420 | 0.17578 | 0.18066 | 0.19366 | 11.448 |
| Banking system z-scores | 0.06751 | 0.14156 | 0.28909 | 0.00036 | 11.913 |
| General government debt | 0.18755 | 0.20567 | 0.03208 | 0.02972 | 12.1253 |
| Capital investments | 0.35795 | 0.00053 | 0.16497 | 0.00247 | 12.544 |

Source: own results

**Table 5.**
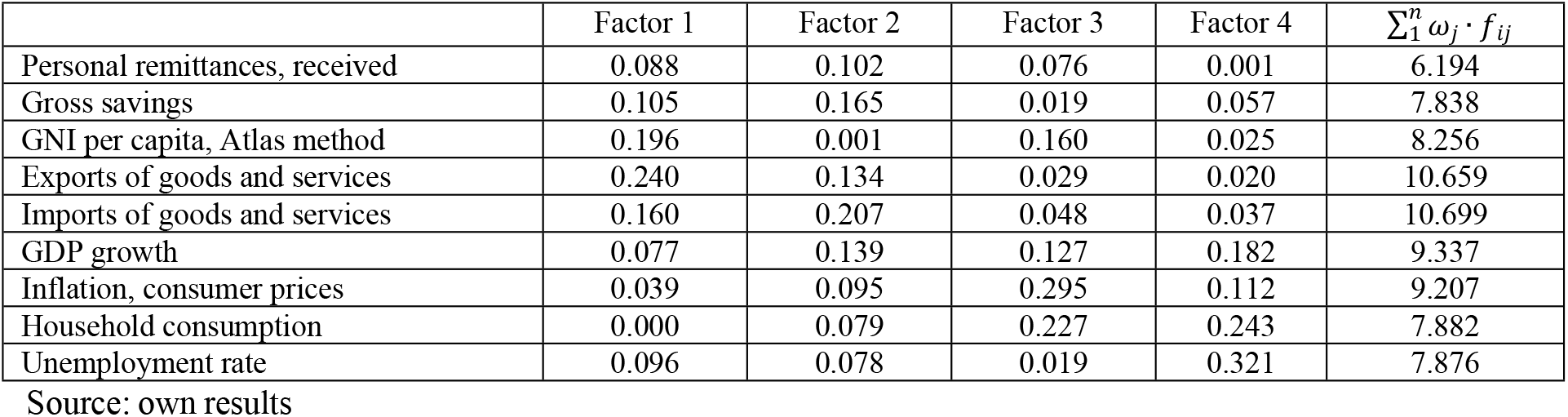
Contribution of economic variables to each factor

**Table 6.** The contribution of political and institutional variables to each factor

| | Factor 1 | Factor 2 | $\sum_1^n \omega_j \cdot f_{ij}$ |
| --- | --- | --- | --- |
| Property Rights Index, IPRI | 0.1488 | 0.0136 | 9.267 |
| Government effectiveness | 0.1973 | 0.0001 | 12.003 |
| Political stability | 0.1806 | 0.0290 | 11.442 |
| Voice and accountability | 0.2023 | 0.0218 | 12.646 |
| Democratic performance numeric | 0.1976 | 0.0313 | 12.512 |
| Corruption Perceptions Index | 0.0730 | 0.9039 | 18.616 |

Source: own results

In order to check the inclusion/exclusion of the indicator in each direction in further research, a selection was made under the conditions of fulfillment (1):

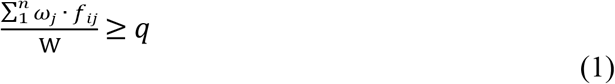

where:

n– the number of factors;

ω_j_ – the percentage of providing variation due to the j-th factor;

fi_j_ – the weight of the i-th variable in terms of the j-th factor;

W – cumulative variation;

q– the critical importance of the relevance of indicators for assessing the manifestation of the consequences of COVID-19 (for financial-budgetary and political-institutional ones is 0.13, for economic – 0.11).

The analysis of the contribution of each change to the variation of the significant factors of the study of the financial and budgetary development of countries allows for sifting out two determinants: General government debt (% of GDP) and Banking system z-scores, index points. The following determinants will be included in the further study: Bank non-performing loans to total gross loans (%), Commercial bank branches (per 100,000 adults), Total reserves (includes gold, current US$), Bank capital to assets ratio (%), Capital investments (% of GDP). Applying a similar methodology to economic determinants, a set of six indicators was obtained: Exports of goods and services (% of GDP), Imports of goods and services (% of GDP), GDP growth (annual %), Inflation, consumer prices (annual %), Household consumption, billion U.S. dollars, Unemployment rate, %. The following indicators were selected for the study of political and institutional development: Government effectiveness, Political stability, Voice and accountability, Democratic performance numeric and Corruption Perceptions Index.

### 4.2. Construction of integral indicators of manifestation of the consequences of COVID-19

Data were normalized to provide a statistical base for the study. For the indicators that are destimulators (in terms of economic determinants: Unemployment rate and Inflation, consumer prices) – Savage normalization (2), for the rest of the indicators – stimulants through natural normalization (3).

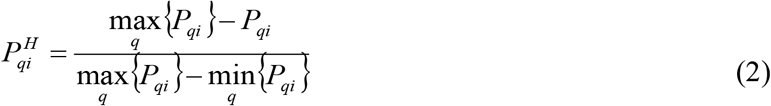

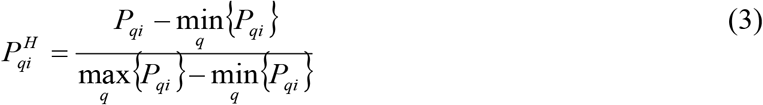

where:

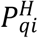 – normalized values by q-year for i-country;

*P*_*qi*_– actual values of the q-year for the i-th country;

min{*P*_*qi*_} – minimum value for q-year for i-country;

max {*P*_*qi*_} – the maximum value for q-year for country i, *q* = 2017..2020, *i* = 1..59.

To build integral indicators of the manifestation of the consequences of COVID-19 for 2017-2020, we will apply the method of group accounting of arguments – convolution of indicators, using the Kolgomorov-Gabor polynomial, which combines additive and multiplicative methods (4):

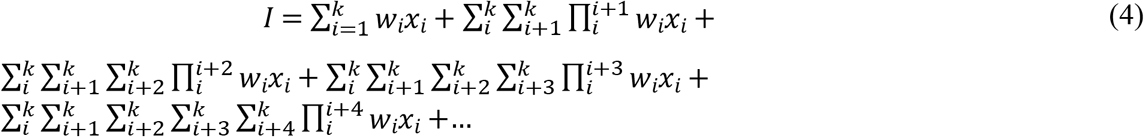

where:

*w*_*i*_ – weights of the i-th factor, we take all weights as one;

*x*_*i*_^*^ – normalized value of the i-th factor.

The normalized results of the integral indices for the assessment of the economic, political-institutional and financial-budgetary development of the countries of the world are presented in Table 7

**Table 7.** Normalized results of integral indices for assessing the economic, political-institutional and financial-budgetary development of the countries of the world

|  | Financial and budgetary |  |  |  | Institutional and political |  |  |  | Economic |  |  |  |
| --- | --- | --- | --- | --- | --- | --- | --- | --- | --- | --- | --- | --- |
|  | 2017 | 2018 | 2019 | 2020 | 2017 | 2018 | 2019 | 2020 | 2017 | 2018 | 2019 | 2020 |
| Austria | 11% | 9% | 11% | 12% | 72% | 78% | 80% | 81% | 16% | 19% | 21% | 19% |
| Belgium | 24% | 21% | 22% | 24% | 70% | 70% | 76% | 77% | 24% | 29% | 28% | 22% |
| Bosnia and Herzegovina | 54% | 44% | 48% | 46% | 55% | 57% | 65% | 62% | 30% | 36% | 41% | 36% |
| Brazil | 14% | 11% | 18% | 20% | 8% | 7% | 9% | 7% | 13% | 13% | 16% | 9% |
| Canada | 7% | 4% | 7% | 10% | 13% | 13% | 15% | 16% | 8% | 6% | 8% | 3% |
| Switzerland | 44% | 40% | 48% | 58% | 86% | 84% | 87% | 89% | 22% | 21% | 23% | 14% |
| Chile | 10% | 8% | 10% | 8% | 93% | 97% | 100% | 100% | 26% | 35% | 30% | 35% |
| China | 100% | 100% | 100% | 100% | 46% | 49% | 50% | 51% | 13% | 20% | 14% | 7% |
| Cyprus | 68% | 38% | 45% | 40% | 4% | 5% | 4% | 5% | 39% | 45% | 42% | 32% |
| Czech Republic | 18% | 16% | 20% | 23% | 40% | 42% | 49% | 44% | 40% | 48% | 52% | 35% |
| Germany | 5% | 3% | 6% | 8% | 45% | 46% | 49% | 50% | 45% | 45% | 43% | 31% |
| Denmark | 11% | 9% | 13% | 15% | 71% | 73% | 62% | 62% | 29% | 26% | 29% | 29% |
| Spain | 32% | 29% | 33% | 31% | 87% | 93% | 98% | 98% | 26% | 27% | 31% | 29% |
| Estonia | 23% | 19% | 18% | 20% | 41% | 43% | 29% | 28% | 12% | 10% | 14% | 2% |
| Ethiopia | 23% | 16% | 14% | 13% | 53% | 57% | 62% | 66% | 42% | 41% | 45% | 37% |
| Finland | 9% | 7% | 2% | 5% | 0% | 1% | 1% | 1% | 22% | 13% | 16% | 3% |
| France | 23% | 20% | 25% | 28% | 91% | 93% | 64% | 67% | 21% | 16% | 21% | 19% |
| United Kingdom | 6% | 4% | 8% | 11% | 50% | 53% | 67% | 64% | 19% | 17% | 21% | 13% |
| Georgia | 46% | 41% | 39% | 34% | 65% | 63% | 71% | 69% | 21% | 22% | 26% | 16% |
| Greece | 48% | 37% | 45% | 38% | 20% | 21% | 19% | 19% | 19% | 26% | 28% | 8% |
| Honduras | 25% | 23% | 22% | 19% | 26% | 26% | 30% | 33% | 6% | 5% | 10% | 3% |
| Croatia | 54% | 45% | 46% | 50% | 6% | 6% | 5% | 5% | 30% | 29% | 26% | 10% |
| Hungary | 46% | 43% | 59% | 57% | 29% | 28% | 32% | 32% | 22% | 24% | 33% | 18% |
| Indonesia | 54% | 48% | 51% | 49% | 26% | 27% | 42% | 45% | 43% | 57% | 52% | 35% |
| India | 34% | 33% | 37% | 40% | 13% | 14% | 19% | 19% | 22% | 25% | 28% | 17% |
| Ireland | 68% | 43% | 77% | 52% | 15% | 16% | 28% | 27% | 28% | 30% | 25% | 6% |
| Iceland | 58% | 43% | 47% | 46% | 70% | 76% | 45% | 49% | 80% | 100% | 82% | 81% |
| Israel | 10% | 8% | 11% | 14% | 81% | 79% | 70% | 69% | 30% | 36% | 27% | 15% |
| Italy | 31% | 20% | 26% | 27% | 33% | 32% | 43% | 40% | 26% | 27% | 29% | 21% |
| Japan | 36% | 35% | 46% | 48% | 34% | 34% | 41% | 44% | 14% | 11% | 14% | 11% |
| Kazakhstan | 26% | 17% | 27% | 31% | 62% | 65% | 52% | 56% | 20% | 18% | 20% | 22% |
| Lithuania | 10% | 7% | 0% | 0% | 4% | 3% | 7% | 8% | 17% | 20% | 26% | 11% |
| Luxembourg | 35% | 26% | 33% | 38% | 43% | 44% | 32% | 32% | 34% | 41% | 45% | 33% |
| Latvia | 23% | 23% | 16% | 15% | 84% | 88% | 79% | 80% | 63% | 87% | 100% | 100% |
| Moldova | 79% | 71% | 51% | 51% | 38% | 40% | 34% | 32% | 26% | 34% | 30% | 27% |
| North Macedonia | 46% | 39% | 44% | 41% | 9% | 9% | 11% | 12% | 25% | 32% | 28% | 16% |
| Malta | 24% | 17% | 23% | 28% | 12% | 14% | 28% | 30% | 10% | 40% | 23% | 9% |
| Montenegro | 50% | 48% | 54% | 57% | 50% | 50% | 52% | 53% | 100% | 95% | 94% | 64% |
| Malaysia | 21% | 15% | 16% | 17% | 16% | 17% | 20% | 19% | 21% | 23% | 23% | 1% |
| Nigeria | 0% | 6% | 9% | 14% | 22% | 26% | 32% | 32% | 41% | 50% | 48% | 32% |
| Netherlands | 2% | 0% | 3% | 5% | 0% | 0% | 0% | 0% | 0% | 1% | 6% | 0% |
| Norway | 33% | 27% | 36% | 35% | 82% | 83% | 88% | 91% | 37% | 42% | 39% | 38% |
| Panama | 57% | 52% | 48% | 30% | 100% | 100% | 69% | 73% | 20% | 16% | 20% | 23% |
| Peru | 17% | 13% | 17% | 13% | 20% | 20% | 23% | 23% | 35% | 33% | 32% | 3% |
| Philippines | 16% | 17% | 20% | 16% | 14% | 13% | 16% | 15% | 17% | 25% | 21% | 7% |
| Poland | 25% | 22% | 26% | 25% | 9% | 11% | 14% | 15% | 35% | 35% | 41% | 13% |
| Portugal | 26% | 18% | 23% | 25% | 32% | 34% | 28% | 28% | 34% | 44% | 43% | 29% |
| Romania | 26% | 21% | 28% | 31% | 56% | 56% | 59% | 57% | 23% | 25% | 28% | 15% |
| Russian Federation | 47% | 37% | 47% | 51% | 20% | 20% | 30% | 31% | 39% | 30% | 32% | 21% |
| Serbia | 63% | 62% | 73% | 72% | 2% | 2% | 6% | 6% | 15% | 19% | 18% | 15% |
| Slovak Republic | 28% | 23% | 27% | 24% | 16% | 16% | 20% | 19% | 16% | 25% | 30% | 24% |
| Slovenia | 24% | 25% | 26% | 24% | 35% | 34% | 53% | 52% | 40% | 51% | 44% | 36% |
| Sweden | 9% | 7% | 9% | 11% | 46% | 47% | 47% | 47% | 44% | 50% | 46% | 37% |
| Thailand | 20% | 20% | 23% | 27% | 85% | 89% | 95% | 95% | 20% | 21% | 24% | 20% |
| Turkey | 34% | 29% | 30% | 37% | 7% | 6% | 13% | 12% | 40% | 48% | 38% | 32% |
| Tanzania | 38% | 37% | 38% | 44% | 4% | 5% | 5% | 6% | 13% | 0% | 0% | 1% |
| Ukraine | 52% | 38% | 48% | 29% | 6% | 6% | 7% | 7% | 23% | 25% | 30% | 19% |
| United States | 36% | 32% | 40% | 42% | 4% | 5% | 8% | 9% | 9% | 13% | 19% | 14% |

Source: own results

### 4.3. Probit/logit modeling of the manifestation of the consequences of the COVID-19 pandemic

In order to identify fluctuations in economic, financial-budgetary and political-institutional development, a binary response model was used: logistic and probit regression. The happiness index of the countries of the world for 2018-2020 (World Happiness, 2022) was chosen as the dependent variable, which consists of a large number of indicators, but the results of the Gallup global sociological survey make up the most significant specific weight in it. The value of the happiness index is in the range from 2.3 to 8, therefore, to measure the fluctuations of this index as a result of the pandemic, the statistical data was coded into a binary system according to rule (5). Normalized composite estimates of the economic, political-institutional, and financial-budgetary development of the world for 2019 – before the start of the pandemic, and for 2020 – the first year of the pandemic, were chosen as independent variables.

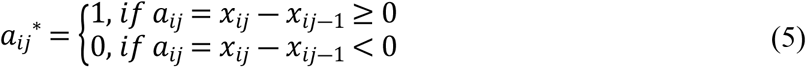

*x*_*ij*_ – normalized values of the happiness index of the ith country of the world, *j* = 2018,…,2020.

In order to build a qualitative and adequate model, logit-(6) and probit-(7) models were built in the study using the Statistica Portable application program package using the Advanced NonLinear Models - Nonlinear Estimation toolkit. However, the results of the constructed probit regression do not satisfy the adequacy criteria, so only the results of the logit regression were included in the further study.

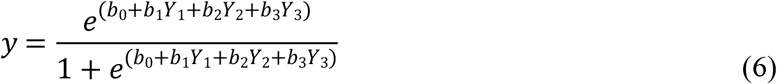

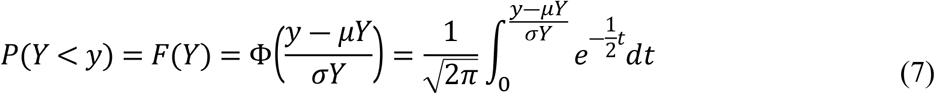

where:

y – variable value Y;

μ*Y* – mathematical expectation Y;

*σY* ― root mean square deviation Y;

*b*_0_ - free member;

*b*_1_ ― coefficient of financial and budgetary development;

*b*_2_ ― coefficient of political and institutional development;

*b*_3_ ― coefficient of economic development.

The results of the non-linear evaluation of the level of happiness depending on the economic, political-institutional and financial-budgetary development for 2019 – before the start of the pandemic, and for 2020 – taking into account the consequences of the pandemic are shown in Table 8, Figure 2, 3 and approximated by equations (8) and (9) for 2019 and 2020, respectively.

**Table 8:** Results of the logit model for assessing fluctuations in the economic, financial, budgetary and political-institutional development of countries as a result of the pandemic

| 2019 year,<br>Loss: Max likelihood Final loss: 34,277611281 Chi-square =8,2677 p=0,04081 |  |  |  |  |
| --- | --- | --- | --- | --- |
| | $b_0$ | $b_1$ | $b_2$ | $b_3$ |
| **Rating** | -0.004254 | -2.55817 | -0.764682 | 5.8668 |
| Odds ratio (units) | 0.995755 | 0.07745 | 0.465482 | 353.1013 |
| Odds ratio (range) |  | 0.07745 | 0.465482 | 353.1013 |
| 2020 year,<br>Loss: Max likelihood Final loss: 23,177167970 Chi-square=13,243 p=0,00414 |  |  |  |  |
| **Rating** | 1.060722 | 2.41581 | -4.27599 | 8.647 |
| Odds ratio (units) | 2.888455 | 11.19886 | 0.01390 | 5694.792 |
| Odds ratio (range) |  | 11.19886 | 0.01390 | 5694.792 |

Source: own results

**Figure 2.**
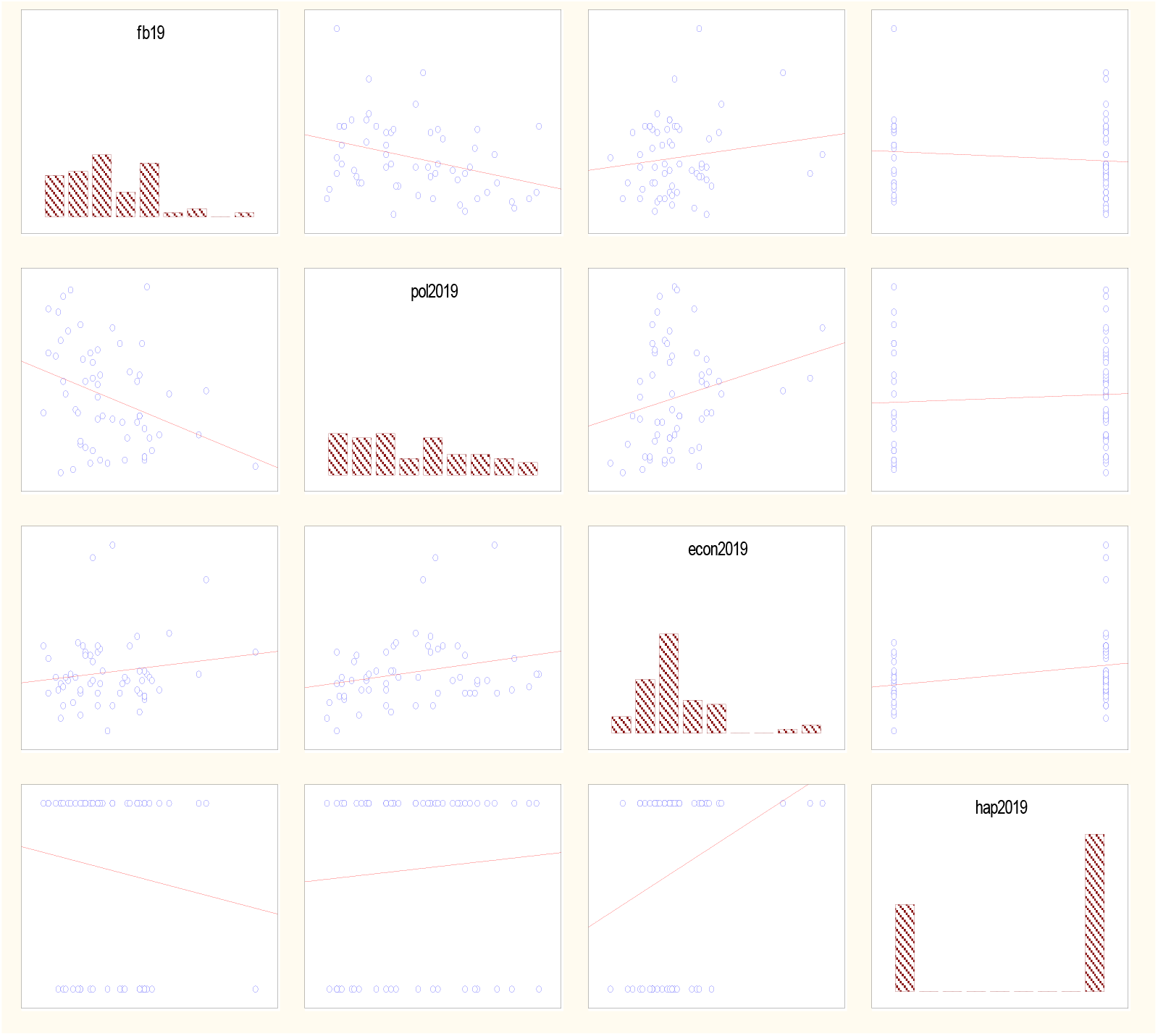
Results of the logit model for assessing the economic, financial, budgetary and political-institutional development as a result of the pandemic Source: own results

**Figure 3.**
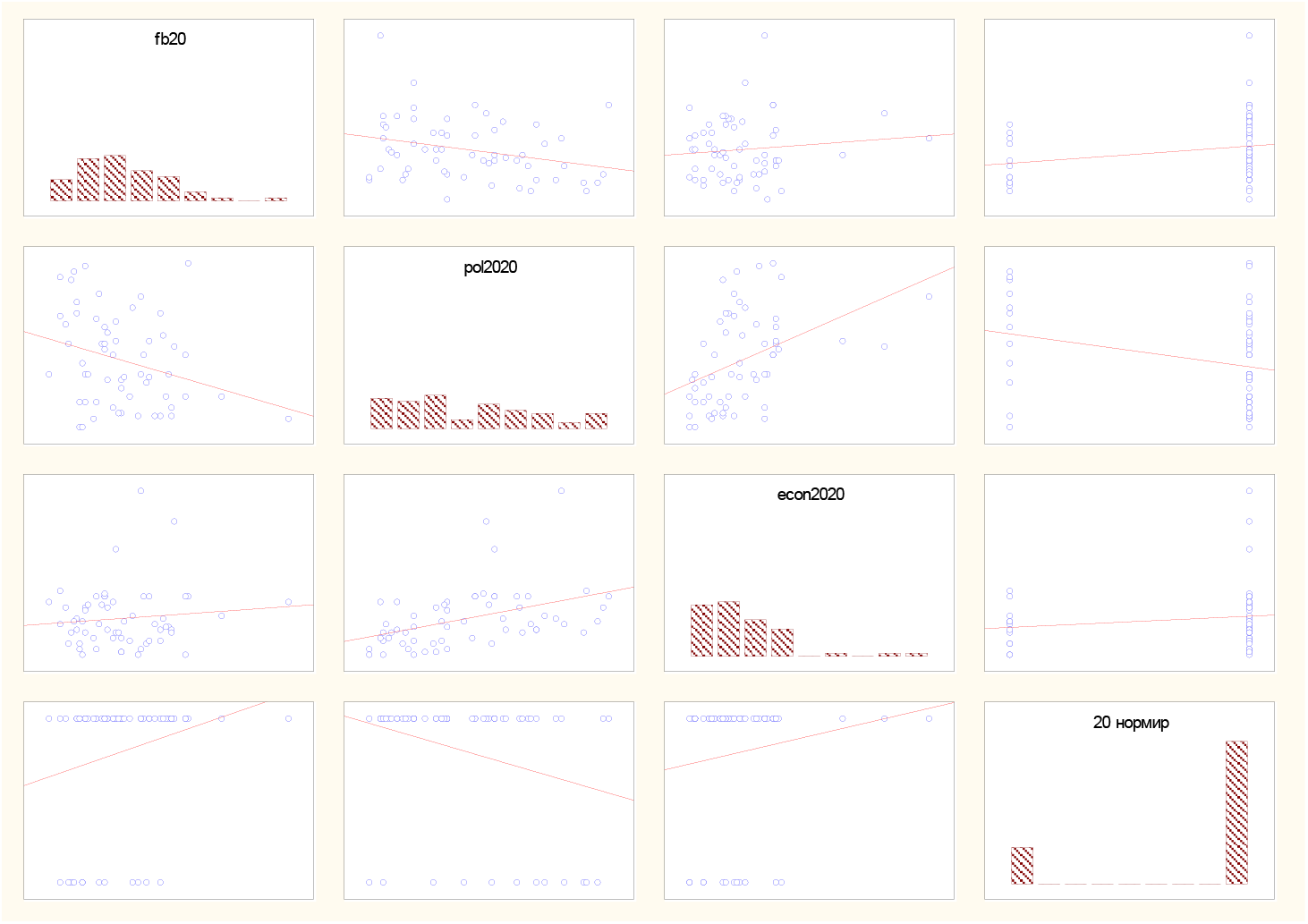
The results of building a logit model for assessing the economic, financial, budgetary and political-institutional development of countries as a result of the pandemic, 2020 Source: own results

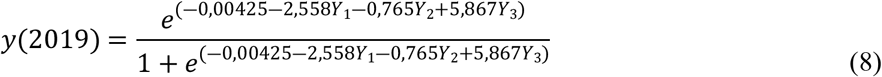

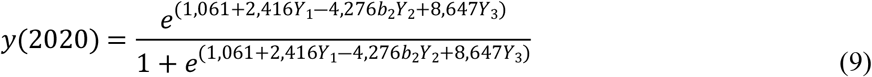

Figures 2 and 3 illustrate the graphical representation of the model building results in 2019 (Figure 2) and 2020 (Figure 3).

The analysis of the significance level for both logit models does not exceed the critical value (0.05) and the chi-square value is also sufficiently large, so the constructed models are adequate. The percentage of correctly predicted results (if the theoretical value is less than 0.5, it is considered 0, if it is more, then 1) for model (8) was 68%, for (9) −80%, which also indicates a high level of correctly guessed results. A comparison of the results of 2019 – before the pandemic and 2020 – during the pandemic allows us to conclude that there has indeed been a transformation of the influence of economic, political-institutional and financial-budgetary development on the general state of “feeling of happiness” among the population. For example, the coefficient of financial and budgetary development in 2019 was 2.558, and in 2020 it became 2.416, so the influence in this direction has increased significantly. The coefficient of economic growth in 2019 was 5.867, and in 2020 it became 8.647, indicating an increase in influence. The political and institutional development coefficient in 2019 was −0.765, and in 2020 it became −4.276. The analysis of changes in the influence on the happiness index of the population of the countries of the world indicates an increase in the influence of the economic and financial-budgetary component and a decrease in the influence of the political-institutional component.

## 5. Conclusions

Overall, in this paper we formed a set of integral indicators indicating the consequences of COVID-19. In addition, we measured the fluctuations in the development of the countries of the world due to the pandemic (through a combination of additive multiplicative convolutions and the Kolmogorov-Gabor polynomial and logit and probit modeling). Relevant economic, financial-budgetary, and institutional-political determinants, which can cause the lack of resilience of national development, have also been identified. Our approach is a fundamentally new, substantive basis for verifying the main channels through which COVID-19 affects the development of countries worldwide. The predictive logit model for assessing economic, financial-budgetary and political-institutional development forms the basis for forecasting the degree of influence of health risks on the development of individual countries and entire regions.

It is noteworthy that when determining the fluctuations of economic, financial-budgetary and political-institutional development, the happiness index was chosen as a dependent variable indicator of well-being in the analyzed countries. The construction of probit regression results showed that the model does not meet the criterion of adequacy, so further calculations to fulfill the research objectives were based only on the logit regression structure. The analysis of the built model made it possible to conclude that economic and financial-budgetary components had a significantly increased influence on well-being in the countries of the world during the COVID-19 pandemic. In contrast, a decrease in the impact of political and institutional indicators was observed, which is vital to take into account in the conditions of further scientific intelligence within the framework of determining the reasons for the non-resilience of national policy to challenges to public health.

## Data Availability

The data can be found at: statista.com, theglobaleconomy.com, and ec.europa.eu

https://ec.europa.eu/

https://www.theglobaleconomy.com/

https://www.statista.com/

## Acknowledgments

This work was supported by the Ministry of Education and Science of Ukraine (research topics No. 0122U000781 “The impact of COVID-19 on the transformation of the medical and social security system: economic, financial and budgetary, institutional and political determinants” and No. 0122U000778 “Socio-economic recovery after COVID-19: modeling the consequences for macroeconomic stability, national security and community resilience”.

